# Research Protocol: Discrete choice modelling to understand the influence of sharing polygenic risk scores related to cardiovascular disease risk with primary care patients

**DOI:** 10.1101/2024.10.18.24315590

**Authors:** Lin Bowker-Lonnecker, Padraig Dixon, Stavros Petrou, John Buckell

## Abstract

**Introduction:** Cardiovascular disease (CVD) is a leading cause of death in the UK and globally. People identified as being at high risk may receive further investigations or preventive treatment. Polygenic risk scores (PRSs) give a summary of overall underlying genetic risk, and may be used to give additional information that GPs can use alongside other information about the patient to determine which interventions, if any, would be beneficial.

**Methods and Analysis:** Two discrete choice experiments (DCEs) with 2000 participants recruited from the UK general adult population. The first DCE aims to determine people’s attitudes about getting their PRS in the context of cardiovascular disease, and what factors may influence this. The second DCE aims to determine how people are likely to react to this risk information, and their stated probability of undergoing further investigation or interventions for disease management. This aims to provide new, quantitative information of whether individuals’ health-related behaviour is likely to be modified by knowledge of one’s PRS. Results from the pilot study will be used to inform the design of the main study, and the analysis will use multinomial logit models. Marginal rates of substitution between attributes, and heterogeneity analysis comparing people with different demographic characteristics, will also be carried out.

**Ethics and Dissemination:** Ethics approval (reference: R89898/RE001) was obtained through the Medical Sciences Interdivisional Research Ethical Committee (MS IDREC) at the University of Oxford. The results of this research will be submitted to academic journals and will be presented at conferences.

## Introduction and Aims

Cardiovascular disease (CVD) is amongst the leading causes of death globally, with approximately 17.9 million people dying of CVD in 2019, which accounts for 32% of global deaths (1). Around 152,000 people died of CVD in the UK in 2016, making it the second most common cause of death, including causing 21% of deaths for those under the age of 75 years. Death rates attributable to CVD vary with factors such as age, region and socioeconomic status, with men being more likely to die of CVD compared to women (2). The British Heart Foundation estimates that CVD costs the UK healthcare system around £10 billion annually, and the wider UK economy around £25 billion annually (3).

Both genetic and lifestyle factors play a role in the development of CVD. Poor diet and nutrition, irregular exercise, and low cardiorespiratory fitness all increase the risk of CVD, and certain conditions such as obesity and diabetes also increase the risk (4,5). Those with family history of CVD are also more likely to develop CVD themselves, with a 60%-75% increased risk if a parent has premature CVD, and an approximately 40% increased risk if a sibling is affected (6).

Polygenic risk influences susceptibility to many diseases, including CVDs such as coronary artery disease (7). There is therefore increasing interest in the potential of polygenic risk scores (PRSs) – aggregated summaries indicating genetic liability to disease-related outcomes – to identify patients with sufficient risk that they may benefit from preventative treatment (8). This could involve more precise identification of those at the highest absolute risk for incident disease, the prediction of progression, or other markers of worse patient outcomes. Currently there is no large-scale implementation of PRSs. Instead, National Health Service (NHS) genetic testing is offered if it is believed the person may have certain genetic health conditions that are typically monogenic, associated with cancer, and/or rare (9). If healthy people want to find out their genetic information (for both ancestry and health reasons), this can be done through a commercial at home direct to consumer DNA test (e.g. 23andMe), and as of 2019 more than 26 million people worldwide had done so using the four leading companies (10). They may also participate in initiatives such as Our Future Health, which plans to calculate PRSs for 5 million study participants (11).

There are number of challenges and concerns associated with the wider use of PRSs. Existing PRS data is primarily from populations of European descent, so it may not be accurate for people in other regions globally nor those in minority ethnic groups within the UK, and there is the risk that the data itself may be used to discriminate against those at higher risk (e.g. through higher insurance premiums) (12). Beyond this, there are privacy and data protection concerns. This is highly personal data, and there needs to be careful consideration of who has access to it and what it can be used for (13). There is often difficulty in interpreting PRSs, and it is important to communicate clearly what the PRS information means, e.g. it is information about some component of underlying risk, but something that alone cannot be used to make a diagnosis. Potential responses to knowledge of PRS may also complex. Having a high PRS could potentially cause stress, however a low PRS may lead people to risk compensate by reducing preventative measures, e.g. not attending screening appointments or maintaining unhealthy behaviours (12).

A 2022 systematic review identified 29 studies relating to predictive models for CVD using genetic risk scores and found that 27 of the 29 studies associated genetic risk scores with the incidence of CVD (14). The results of 23 studies indicated some improvement in clinical utility (compared to traditional models of predicting CVD, either through improved discrimination or improved risk classification. However, effects were modest, didn’t necessarily reflect pragmatic clinical implementations or cost-effectiveness, the methods used were heterogeneous and the authors noted that further research is needed (14). A 2023 scoping review with a broader focus on PRSs for cardiometabolic diseases, with 82 studies meeting the inclusion criteria, concluded that predictive accuracy was improved by integrating PRSs with traditional clinical risk tools (15). However, the extent of such improvements was modest and doesn’t in itself constitute evidence for or against the use of PRSs in the prediction or management of CVD.

The 2023 scoping review also noted that there is limited research that exists on investigating whether providing PRS information to people would lead them to improving health behaviours such as increasing levels of exercise or eating a healthier diet (15). In an observational follow-up study that examined PRSs, 7342 Finnish adults were provided with PRSs using an interactive web tool. They found that 89% of participants said that getting this risk information motivated them to take better care of their health, with 42.6% of those with high risk of atherosclerotic CVD making one or more health behavioural changes after 1.5 years, compared to 33.5% of those at a lower risk. This may on average correspond to a 10-15% relative reduction in their risk of CVD (16). The MI-GENES clinical trial randomized participants with intermediate risk of coronary heart disease and gave some their Framingham risk score, and others their integrated risk score (IRS), which included their Framingham risk score and their PRS. A recent preprint suggests that participants who received an IRS had a lower incidence of major adverse cardiovascular events, and a higher statin therapy initiation rate and duration, after 10 years of follow-up compared to those who only received their Framingham risk score (17). A life insurance company, MassMutual, recently collaborated with Genomics PLC to offer genetic risk assessment services to 1400 policyowners. Of these, 1 in 5 learned that they were at higher risk of some form of preventable disease, and 71% of respondents reported that planned to take preventative action. Amongst this group, if a high-risk result was received, health care visits doubled for the four months after the test (18). This was according to a commercial press release, but can be seen as an example of where genetic risk assessments have been implemented and used to make decisions.

The aim of this research is to provide additional quantitative information on whether individuals’ health-related behaviour is likely to be modified by knowledge of one’s PRS in relation to cardiovascular disease, and on whether people would be willing to get their PRS in this context.

## Methods and Analysis

### Experimental Design and Methods

#### Discrete Choice Experiments

A discrete choice experiment (DCE) is a quantitative tool seeking to measure the preferences of participants. DCEs are increasingly being used in healthcare (19), and in recent years DCEs have been used more in health-related studies than in other fields (20). In a DCE, participants are asked to select their preferred choice out of a limited number of options, where each option is described by a set of attributes (e.g. cost) and their levels (e.g. higher or lower cost).

DCEs can quantitively assess these preferences, and model the influence of specific attributes on choices. They can also be used to model scenarios that currently do not exist. However, they are a stated preference method and subject to hypothetical bias, and do not capture longitudinal preferences. Attribute selection is very important, as DCE modelling can only show associations between listed attributes and not others (21), and key attributes being missed leads to omitted variable bias (22).

DCEs have been used to measure PRS-related behaviours. A search of Medline using search terms relating to DCEs and PRSs found three papers that explicitly use DCEs relating to PRSs: one on how surgeons react to polygenic risk information for diverticulitis (23), one on how the UK general public perceives using PRSs in cancer screening compared to using other criteria (24), and one on the preferences of the Australian general public on different aspects of a PRS test to estimate cancer risk (25). A 2024 systematic review summarises DCEs involving preferences for genetic testing to predict the risk of developing hereditary cancers (26); however, only one study (25) within the systematic review mentioned PRSs explicitly. There do not appear to be any currently published DCEs relating to PRSs for CVD.

#### Eligibility Criteria and Identification of Participants

For this DCE, the following eligibility criteria will be used:

- Adults (18 years or older)
- Able to read English
- Willing and able to give informed consent for the participation of the study

Participants need to have access to a device (computer, tablet, smartphone) and the internet to complete the survey.

A total of 2,000 study participants will be recruited by a survey company, SurveyEngine GmbH, through email lists to people who have consented to be contacted for this purpose. Of these 2000 people, 50 people will take part in the pilot study.

Quotas will align distributions of participant gender (male, female), age (age bands: 18-34, 35-49, 50-64, 65+), ethnicity (white and non-white) and region (north, midlands, south, London) with those of the overall general population as per the 2021 census, with the aim of getting a study population that is more representative to the overall population of England than without quotas. This will be large enough to perform heterogeneity analysis based on various demographic variables. Informed consent will be obtained from the participants at the start of the survey, and participants can withdraw consent at any time by exiting the survey.

#### Additional Demographic Questions

Additional demographic questions will be asked of the participants, which will be used in the heterogeneity analysis. These questions can be grouped into four broad categories: health information (such as existing medical conditions), demographics (such as gender and ethnicity), existing attitudes (such as on data privacy), and theoretical willingness to make behavioural changes (such as stated willingness to exercise more if given new risk information).

#### Attribute Selection

To determine potential attributes and questions to ask in the DCE, a brief background search of relevant literature was conducted using the Medline (Ovid) database. This searched for attribute selection methodology, on DCEs focusing on CVD, and on DCEs focusing on PRSs (also mentioned in the ‘Discrete Choice Experiments’ section of this protocol paper). An initial list of attributes was created, which was then discussed with collaborators and further narrowed down. After creating the long list, patient and public involvement (PPI) was received to ensure potential attributes had not been missed, and to determine if any attributes or demographic questions required modification. From this, it was determined that there would be two different sets of DCEs: one on whether getting a PRS would change a person’s health choices, and the other on whether people are willing to get their PRS to begin with.

The potential for bias based on the order in which the DCEs are presented was considered. In reality, getting one’s PRS will not be compulsory, and one would make the choice on whether to get their PRS before one is able to get the information from it. Therefore, the DCE determining whether somebody is willing to get a PRS will be presented first, followed by the DCE about how people respond to the risk information that is required. However, if somebody is against getting their PRS, they may be biased towards selecting options suggesting that they would change their health choices less on PRS information. A heterogeneity analysis can determine whether this is the case. There will be demographic questions before, between and after the DCEs.

Additional PPI was conducted for the final version of the attributes, to ensure that the attributes and levels are understood by participants. Based on this, the wording of several attributes was updated to make it easier to understand, including for the ‘accuracy of test’ and ‘incidental findings’ attributes. The level of the ‘lifestyle support’ attribute was updated from ‘web information’ to ‘web or printed information’, to acknowledge that some people might not have easy access to the internet.

An additional attribute was also added to the first DCE regarding time taken between requesting the test and receiving results. This attribute was initially not included in part because it was not included in the most comparable literature, and in part because the time attribute is less important when determining underlying risk compared to diagnosing acute conditions. However, it is likely that people assume that private healthcare has shorter waiting times, and one PPI participant explicitly thought this. Therefore, this either needs to be mentioned in the scenario and kept constant, or included as an attribute. Depending on the results of the pilot study, this attribute may be included in the main study or removed.

##### Attribute Set 1: Willingness of getting PRS

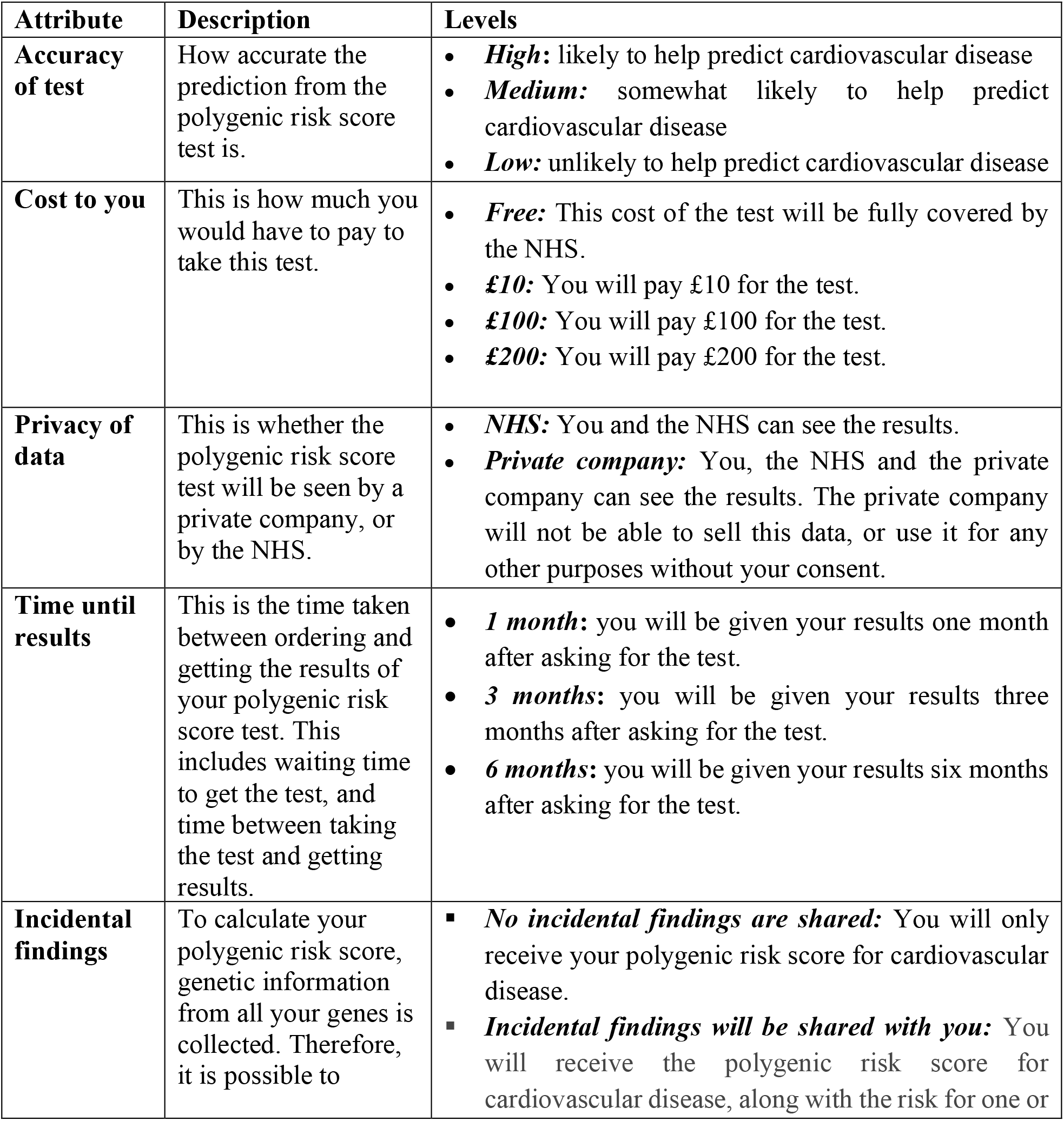
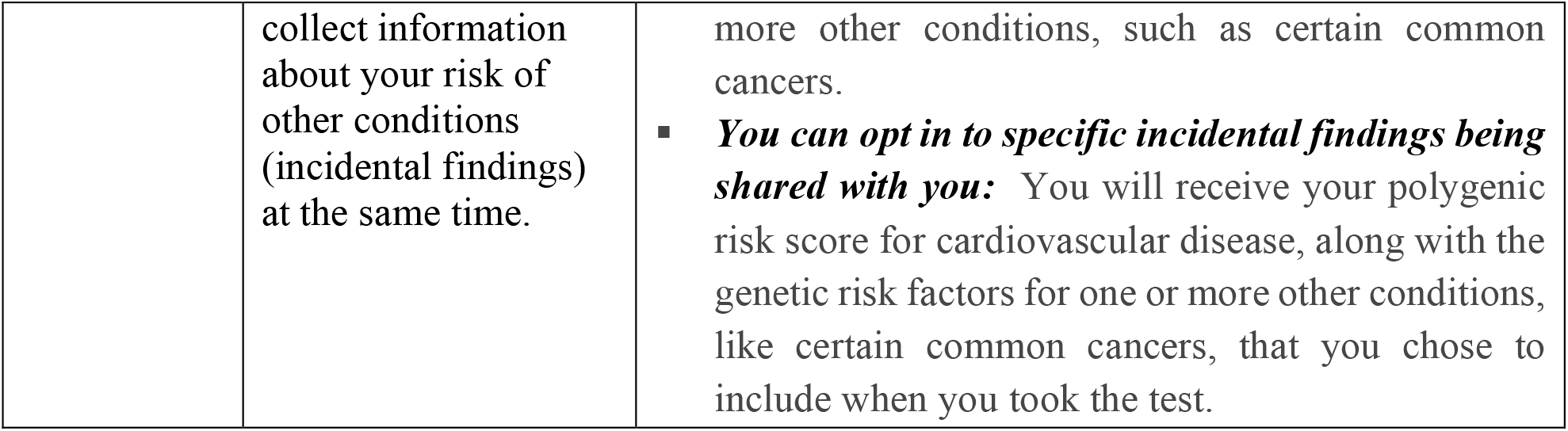

The ‘cost to you’ values are based on the NHS single prescription charge, which is £9.90 at time of writing *(27)*, and the private values are rounded based on 23andMe prices *(28)*. Currently 23andMe estimates a sample processing of 4-6 weeks and a delivery time of 2-4 weeks in the United States *(29)*. While this is unlikely to be representative of larger scale or screening programme use, this has been used to help determine the lower two levels for the ‘Time between asking for test and getting results’ attribute.

To test people’s willingness to get a PRs, an opt out option (i.e. not getting a risk score) should be provided. In the pilot, there will be no forced choice – if people always prefer not to get their PRS, this will also be valuable information about its likely uptake.

For the pilot study, all attributes will be dummy coded, which is a way of coding discrete levels of categorical attributes *(30)*. This includes the cost, as it is possible that willingness to pay is not linear, e.g. people have a strong preference for the tests to be free. If the pilot shows that the willingness to pay is greater than £200, one or both levels corresponding to the private cost of doing the PRS test might be increased.

##### Attribute Set 2: Response to Risk Information

This DCE provides the participant with a hypothetical result to their PRS test. This scenario has the following options:

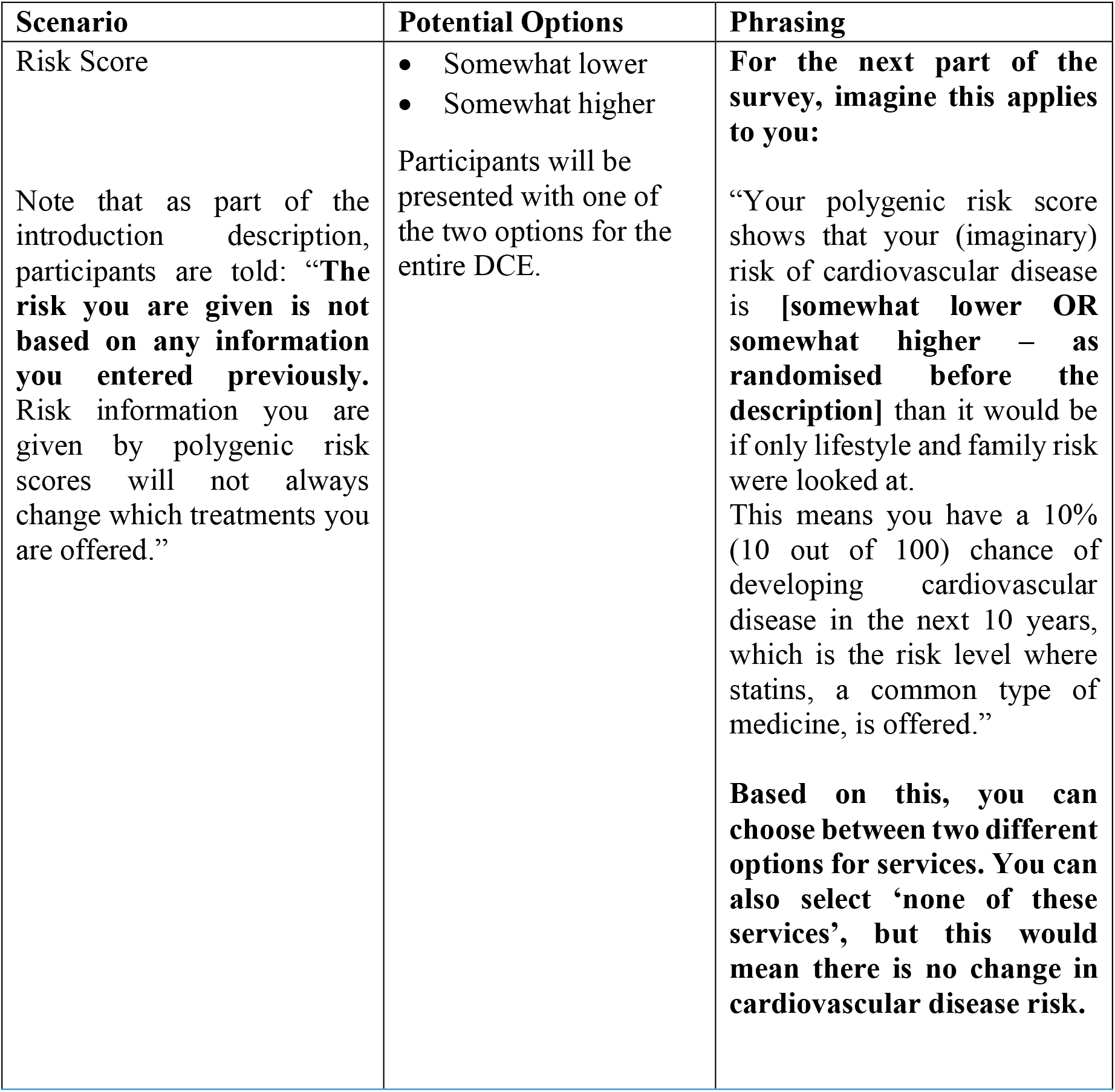

| Scenario | Potential Options | Phrasing |
| --- | --- | --- |
| Risk Score<br>Note that as part of the introduction description, participants are told: **“The risk you are given is not based on any information you entered previously. Risk information you are given by polygenic risk scores will not always change which treatments you are offered.”** | • Somewhat lower<br>• Somewhat higher<br>Participants will be presented with one of the two options for the entire DCE. | **For the next part of the survey, imagine this applies to you:**<br>“Your polygenic risk score shows that your (imaginary) risk of cardiovascular disease is [somewhat lower OR somewhat higher – as randomised before the description] than it would be if only lifestyle and family risk were looked at.<br>This means you have a 10% (10 out of 100) chance of developing cardiovascular disease in the next 10 years, which is the risk level where statins, a common type of medicine, is offered.”<br>**Based on this, you can choose between two different options for services. You can also select ‘none of these services’, but this would mean there is no change in cardiovascular disease risk.** |

After this scenario, the participant will be presented with choice sets containing the following attributes and levels.

| Attribute | Description | Levels |
| --- | --- | --- |
| **Medications** | You may be offered preventative medication options which may include statins. All medications have potential side effects. Medications are free if you are over 65, come at no additional cost if you already have the NHS prepayment certificate, otherwise will cost ~£10 a month or ~£115/year with the NHS prepayment certificate. | • **None:** No preventative medication will be offered in your case.<br>• **One medication (low chance of side effects):** You may have minor side effects, such as headache, dizziness, mild muscle pain, or nausea. These affect 1 out of every 100 people.<br>• **One medication (moderate chance of side effects):** You may have minor side effects, such as headache, dizziness, mild muscle pain, or nausea. These affect 1 out of every 50 people.<br>• **Two medications (moderate chance of side effects):** You may have minor side effects, such as headache, dizziness, mild muscle pain, or nausea. These affect 1 out of every 50 people. |
| **Additional monitoring** | You may be offered more regular monitoring appointments. | • **None:** You will attend your GP as you normally would, otherwise monitoring is by the NHS Health Check (once every 5 years).<br>• **Occasional monitoring:** You will attend occasional monitoring checkups with your GP (once every 2 years)<br>• **Regular monitoring:** You will attend regular monitoring checkups with your GP (once a year) |
| **Lifestyle support** | You may be given additional support to improve lifestyle factors, particularly increasing your amount of exercise and improving your diet. | • **None:** You will attend your GP as you normally would, otherwise monitoring is by the NHS Health Check (once every 5 years).<br>• **Occasional monitoring:** You will attend occasional monitoring checkups with your GP (once every 2 years)<br>• **Regular monitoring:** You will attend regular monitoring checkups with your GP (once a year) |
| **Reduction in risk of heart attack or stroke.** | Based on the above services, your hypothetical (imaginary) cardiovascular disease risk will be reduced by the following amount. | • **Low:** Up to 1% absolute reduction in disease risk. This gives you a remaining risk of 9-10% chance of developing cardiovascular disease in the next 10 years.<br>• **Medium:** Up to 2% absolute reduction in disease risk. This gives you a remaining risk of 8-10% chance of developing cardiovascular disease in the next 10 years.<br>• **High:** Up to 3% absolute reduction in disease risk. This gives you a remaining risk of 7-10% chance of developing cardiovascular disease in the next 10 years. |

All attributes will be dummy coded. It is assumed that either one or two prescribed medications will make it worthwhile for someone to purchase the NHS prepayment certificate, which costs £114.50 for 12 months at time of writing (31), rather than paying the prescription cost each time. The side effects of medications are based on common side effects of statins (32). The change in risk of heart attack or stroke is based on values that seem reasonable for statins (33) or exercise (34).

An opt out option is also provided, in case participants prefer neither option. There will be no forced choice in the pilot, but if many pilot participants choose the opt out option, then participants may additionally be asked for their preference in the main study.

#### Generating Choice Sets

A Bayesian D-efficient design will be used to create the set of choice tasks (35). The aim of a Defficient design is to maximise the statistical efficiency of the design for a given number of choice tasks (35). For the pilot study, the D-efficient design will use uniform distributions (36). Unless the attribute direction is clear a priori (e.g. lower cost is better than higher cost, higher accuracy is better than lower accuracy), zero priors will be used. PPI helped identify attributes where there was uncertainty around the direction of people’s preferences. Choice sets where there is a dominant option (i.e. one option is clearly better than the other, for example better in every way) will not be used (37). These do not give additional information about how people make choices, as people are likely to select the better option in almost all cases.

While the source of the PRS (NHS vs private company) appears to have an obvious cost association (free or £10 for NHS; £100 or £200 for private company), a test that is available for free on the NHS may be analysed by a private company. The fact that the privacy of PRS is ‘private’ is therefore not intrinsically linked to the cost attribute, although the levels for the NHS option will be limited to free or £10. This was raised in the second round of PPI, and sometimes stood out to the PPI contributors. However, when it was made clear that the attribute related to data sharing, they appeared to understand this. The attribute name was therefore revised to ‘Privacy of data’ after this round of PPI to reflect this. However, if this is found to be confusing in the pilot study a limit will be put on these attributes corresponding to the obvious cost associations mentioned above.

#### Data Collection and Data Quality

The main purpose of the pilot study is to analyse the experimental data and produce priors (i.e. parameter estimates) that can be used to update the experimental design and maximise its efficiency for the main data collection. The pilot provides further opportunities to improve the survey and the experimental design (38). Additional questions will be included at the end of the pilot study including about how difficult the survey was to complete and any comments or reflections on the attributes and levels. Therefore, some aspects of attributes and levels provided in this protocol paper may differ from those used in the final survey. Where this occurs, that this has happened and the reason for it will be noted when the results are published.

Steps will be taken in the survey to assess or increase the quality of the data. There will be a question at the end of the survey asking the participant how carefully they answered the questions. The participant will also not be able to skip questions (forced responses) to encourage them to read and answer the questions (although there is a ‘prefer not to answer’ option for many of the demographic questions), and there will also be an overall minimum time for completion of the survey (35). This time limit will be based on timings from the pilot study. After each DCE, the participant will be asked which of the attributes was most important for them when making their choice, which can be checked against the DCE data.

Before the participants start each of the DCEs, a description of each of the attributes and possible levels will be given. Key terms (e.g. ‘polygenic risk score’) will be defined at the start of the experiment, and definitions of key terms will be available to the participant during the DCEs. This will reduce that chance of the participant misunderstanding the question and therefore giving inaccurate responses. One of the additional questions at the end of the pilot study is about whether the participant found any of the questions confusing, which will allow for the survey to be edited for clarity before the main study, if necessary.

To obtain balance (ensuring that all the attribute levels are used an equal number of times) we use 12 choice sets with attributes having 2, 3 or 4 levels. 24 choice sets per participant between 2 DCEs is likely too high, and an upper limit of 18 choice sets per participant has been suggested (35). This means blocking will be used, giving 6 choice sets per participant per DCE, and half the participants see each block of choice sets (35). There will be an additional example choice set before the start of the first DCE, to ensure that the participant understands how the experiment works and what the attributes and levels are. This gives 12 choice sets, and one example choice set, in total per participant.

#### Statistical Analysis

Analysis will be done on R software using the Apollo package. The data will initially be analysed using a multinomial logit (MNL) model. While this is a useful initial model, MNL models have some restrictive assumptions such as independence of irrelevant alternatives (e.g. adding a third option does not change a preference order between two other options presented initially), and assuming identical error distributions across alternatives or across individuals (39). Therefore, further modelling will relax some of these assumptions.

Marginal rates of substitution between attributes will be calculated to determine the willingness to trade the attributes off against each other (39), including on cost against the other attributes for DCE 1 (willingness to get PRS) and change in risk of heart disease or stroke against the other attributes for DCE 2 (response to risk information).

Deterministic heterogeneity analysis will be performed according to individual characteristics (including age), medical characteristics (e.g. existing medical conditions), and attitudes (e.g. concern waiting on medical results). Random heterogeneity can be accommodated using discrete or continuous mixture models.

### Ethics Approval and Dissemination

The Central University Research Ethics Committee (CUREC) is responsible for the ethical review process in the University of Oxford. CUREC 1 approval for lower risk research involving human participants and/or their data was sought, and this was done through the Medical Sciences Interdivisional Research Ethical Committee (MS IDREC). Ethics approval was obtained on 12^th^ December 2023 (reference: R89898/RE001). Ethics amendments were made to reflect changes made after the second round of PPI, and additional feedback after seeing an initial mock-up of the survey.

The results of this research will be submitted to academic journals and will also be presented at conferences. The primary author is a DPhil student at the University of Oxford, and the results will also make up a part of their thesis.

## Data Availability

Not applicable as this is a protocol paper with no primary data.

